# Impact of travel distance to revascularization services on amputations in patients with Peripheral Arterial Disease (PAD): a 10-year analysis of the Public Healthcare System in Brazil Data

**DOI:** 10.64898/2025.12.12.25342148

**Authors:** Júlia Freire Castanheira de Paiva Ferreira, Clara Sanches Bueno, Gabriely Rangel Pereira, Marina Martins Siqueira, Felipe Soares Oliveira Portela, Marcelo Fiorelli Alexandrino da Silva, Antonio Eduardo Zerati, Marcelo Passos Teivelis, Nelson Wolosker

## Abstract

**Background:** Peripheral Artery Disease (PAD) is a progressive condition that can lead to major amputation if not properly managed. Geographic barriers may influence access to vascular care and outcomes.

**Objective:** To assess the impact of geographic distance on amputation rates and to evaluate mortality and hospitalization length among PAD patients treated in Brazil’s public healthcare system.

**Methods:** A nationwide, retrospective, population-based analysis was conducted using DATASUS data from 2015–2024, including 335,716 PAD hospitalizations and 70,602 amputations. Logistic regression models evaluated factors associated with amputation and death.

**Results:** Amputation risk increased among patients treated outside their municipality and in state-managed hospitals. Male sex and older age were associated with higher odds of amputation, whereas women, though less frequently amputated, had greater mortality and longer hospital stays when amputation occurred. Mortality following amputation remained elevated nationwide. Regional variations reflected disparities in access to specialized vascular services.

**Conclusion:** Geographic and structural inequalities increased PAD-related amputations and mortality in Brazil’s public system. Greater distance to care and treatment outside the municipality were key predictors of poor outcomes.

## Introduction

Peripheral Artery Disease (PAD) is a chronic condition characterized by the obstruction of arteries, which impairs blood flow. (1) This can lead to symptoms such as intermittent claudication (IC) and critical limb ischemia. (2,3) If not managed properly, PAD can result in limb amputation. (4) Without treatment, the risk of amputation in PAD patients is approximately 25%. (5) Additionally, patients with PAD have a mortality rate nearly twice that of individuals without the disease over 10 years. (5–7)

Worldwide, approximately 5.56% of individuals over the age of 25 are affected by PAD. (7,8) Delays in starting treatment or extended waiting periods for therapeutic interventions may significantly contribute to disease progression and the likelihood of major amputation. (5,6,9) An 18-year cohort study revealed that approximately 16% of patients underwent amputation without having prior revascularization. (10) Furthermore, population-level analyses conducted over two decades have demonstrated a decline in PAD-related amputations, which coincides with an increase in the use of revascularization procedures. (11)

These findings highlight the significance of structured vascular care pathways in preventing disease progression and improving limb salvage. (5–7,9,11,12) Nonetheless, non-clinical factors such as geographical and logistical barriers have gained recognition as influential in shaping PAD outcomes.

A recent Japanese study identified the distance from a patient’s residence to healthcare facility as an independent predictor of lower-limb amputation, even after adjusting for disease stage and comorbidities. (13) Each incremental increase in distance was significantly associated with a higher risk of amputation, underscoring the need to address disparities in access to specialized care. (13,14) Similarly, a British study found that patients initially admitted to regional hospitals experienced substantial delays in revascularization compared to those treated directly at tertiary referral centers. These delays, often exceeding two weeks, were associated with increased rates of major amputation and in-hospital death. (14)

These observations suggest that both residential location and the structure of healthcare delivery systems may impact the likelihood of limb salvage, making them key considerations in PAD outcome assessments. In addition, a recent investigation into amputations within Brazil’s public healthcare system highlighted a rising trend, identifying PAD as the primary diagnosis linked to major limb loss. (12)

Brazil is a country of continental dimensions, where 71,5% of the population relies exclusively on the public health system (approximately 150 million inhabitants). These patients are referred to specific locations designated by the government.

This study represents the first nationwide analysis in Brazil aimed at evaluating PAD patient outcomes from 2008 to 2024 within the public healthcare system. During these 17 years of observation, 335,716 cases of PAD treatment in hospitals were analyzed, where 70,602 limb amputations were performed. The primary objective of this study was to assess the impact of geographic distance on revascularization facilities on amputation rates. A secondary goal was to evaluate mortality and the length of hospital stay among cases with amputation. This investigation examined the relationship between patient residence location and limb loss, focusing on individuals treated in the public sector.

## Materials and Methods

This was a retrospective, cross-sectional, population-level study conducted between 2015 and 2024. A total of 335,716 PAD treatments were performed during hospital admissions, where a total of 70,602 limb amputations were analyzed. The study was divided into two groups: those who received clinical, open revascularization or plus endovascular repair classified as non-amputation, and the amputation group.

All relevant data were sourced from the public health database and the Tab Net platform of the System of Hospital Information of the Unified Health System (SIH-SUS). (15) This platform systematically records treatment information as mandatory documentation for reimbursement eligibility and provides data on the procedure or treatment during hospitalization.

The study focused on several International Classification of Diseases (ICDs) codes, which were select and extracted for analysis: I70.2 (Atherosclerosis of extremity arteries), I70.8 (Atherosclerosis of other arteries), I70.9 (Generalized and unspecified atherosclerosis), I74.1 (Embolism and thrombosis of other and unspecified parts of the aorta), I74.3 (Embolism and thrombosis of arteries of lower limbs), I74.4 (Embolism and thrombosis of arteries of unspecified limbs), I74.5 (Embolism and thrombosis of iliac artery), I74.8 (Embolism and thrombosis of other arteries) and I74.9 (Embolism and thrombosis of unspecified artery).

The study protocol received approval from the institutional ethics review board committee and was institutionally registered under the number 7.092.654. As the research utilized fully de-identified records, individual consent was not required, in accordance with ethical guidelines for secondary data analysis that protect patient anonymity.

The extracted data included the prevalence of sex (biological assignment at birth), age, geographic region, mortality rates, length of hospital stay, distance travelled, number of amputations, angioplasty, revascularization, and clinical treatments performed in-hospital. The distance travelled was calculated from the zero point of the municipality where the patient resides and the zero point of the corresponding hospital.

Data were described using absolute and relative frequencies for qualitative variables, and using means, standard deviations, medians, quartiles, and observed ranges for quantitative variables. Factors associated with amputation or death were analyzed using binary logistic regression, with results expressed as estimated odds ratios, corresponding confidence intervals, and p-values. Other risk factors could not be included because the data extraction was based on ICD codes rather than on the patient.

Automated dataset extraction was performed using Python (version 2.7.13; Beaverton, OR, USA) in a Windows 10 environment. Data fields were systematically identified and manipulated using Selenium WebDriver (version 3.1.8; Selenium HQ), in combination with the Pandas library (version 2.7.13; Lambda Foundry, Inc., PyData). The complete list of procedural codes used in the data extraction process is available in Appendix 1. Temporal trends in procedures were analyzed through linear regression models, while associations were examined using Spearman’s rank correlation method.

We initially analyzed the Procedures performed, then looked at the Patient and Procedure Characteristics, followed by the Distribution of Procedures by region and year, the Analysis of Procedures and Care Location in Relation to Residence, the Univariate logistic-regression models for amputation by Procedure, Distance, Sex, Region and Hospital Management, the Simple logistic-regression models for mortality among amputation cases, and finally the Simple regression models for length of hospital stay among the amputation cases.

### Statistical analysis

The statistical analysis was performed using SPSS version 20.0 (IBM Corp, Armonk, NY). Generalized linear models with a Gamma distribution were applied to assess differences in procedure rates across regions. For sex and age comparisons, Poisson-distributed generalized models were employed, with the number of procedures included as an offset. Statistical significance was defined as a p-value < 0.001.

## Results

Data from 335,716 cases of PAD treatment in hospital were analyzed. Most procedures were clinical, and amputations accounted for 21% of cases. All procedures performed are presented in Table 1.

**Table 1:**
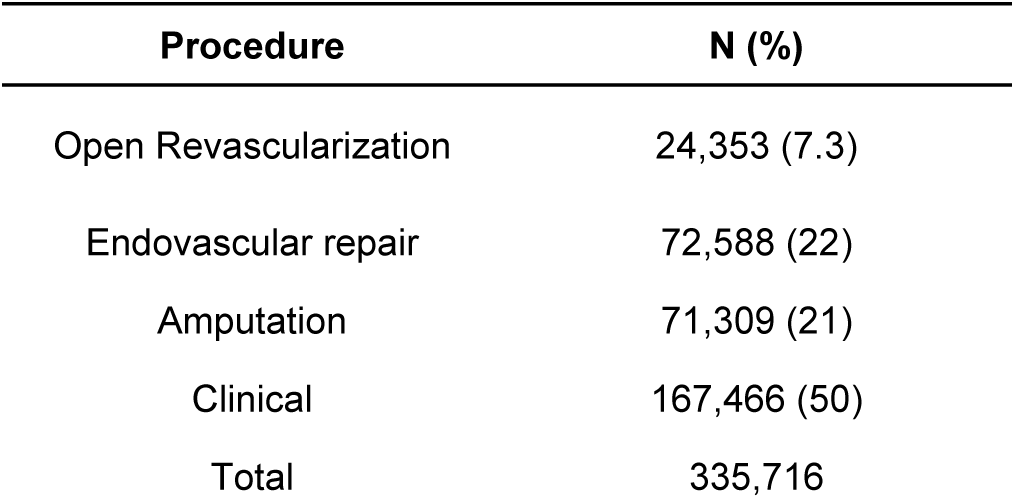
Procedures performed.

| Procedure | N (%) |
| --- | --- |
| Open Revascularization | 24,353 (7.3) |
| Endovascular repair | 72,588 (22) |
| Amputation | 71,309 (21) |
| Clinical | 167,466 (50) |
| Total | 335,716 |

Patient and procedure characteristics from the non-amputation and amputation groups are presented in Table 2. Most patients were male. The average age was 68 years in the groups. ICU admission occurred in 11% of all cases versus 17.7% of amputees; the median ICU stay was 3.0 days. Approximately half of the cases were under complete municipal management. Emergency hospitalizations accounted for more than 84% in both groups.

**Table 2:** Patient and Procedure Characteristics: Non-Amputation vs. Amputation Subgroup.

|  | Non-Amputation<br>N = 264,813<br>N (%) | Amputation<br>N = 70,903<br>N (%) |
| --- | --- | --- |
| Male sex | 145,963 (55.12) | 44,037 (62.11) |
| Average age | 67 [59; 76] | 68 [60; 77] |
| ICU admission | 23,342 (8.81) | 12,536 (17.68) |
| Average days in ICU | 3.0 [1.0; 5.0] | 3.0 [2.0; 7.0] |
| Full municipal management | 136,472 (51.54) | 34,879 (49.19) |
| Urgency hospitalization | 218,752 (82.61) | 62,890 (88.70) |

Distribution of procedures between 2015 and 2024 by region for both groups is presented in Table 3. The amputation rate varied from 19 % to 23 % over the period, being highest in the MidWest region (25.5 %) and lowest in the North (16.8 %). The Southeast region accounted for the largest proportion of cases (46%), with 21% resulting in amputation. Over the study period, the total number of PAD cases increased from 8.4% in 2015 to 11.7% in 2022.

**Table 3:** Distribution of Procedures by region and year.

|  | Non-Amputation<br>N = 335,716* | Amputation<br>N = 70,903** |
| --- | --- | --- |
| Region |  |  |
| Southeast | 121,964 (79.0) | 32,496 (21.0) |
| Midwest | 14,413 (74.5) | 4,926 (25.5) |
| Northwest | 62,306 (77.1) | 18,457 (22.9) |
| North | 6,159 (83.2) | 1,240 (16.8) |
| South | 59,971 (81.3) | 13,784 (18.7) |

| Year |  |  |
| --- | --- | --- |
| 2015 | 22,674 (80.0) | 5,663 (20.0) |
| 2016 | 23,215 (80.1) | 5,781 (19.9) |
| 2017 | 24,669 (80.7) | 5,883 (19.3) |
| 2018 | 26,642 (80.2) | 6,582 (19.8) |
| 2019 | 26,589 (79.4) | 6,887 (20.6) |
| 2020 | 25,089 (76.9) | 7,554 (23.1) |
| 2021 | 26,700 (76.8) | 8,084 (23.2) |
| 2022 | 30,688 (78.0) | 8,659 (22.0) |
| 2023 | 29,844 (78.1) | 8,361 (21.9) |
| 2024 | 28,703 (79.4) | 7,449 (20.6) |

\* Percentage of total cases / \*\* Percentage of amputations by region or year category

The analysis of procedures and care location in relation to residence is presented in Table 4. Procedures performed outside the municipality of residence accounted for roughly half of the cases in both groups. The average travel distance was 5 Km (range 0 to 3,396 km) for all cases and 6 Km (range 0 to 2,906 km) for amputees.

**Table 4:**
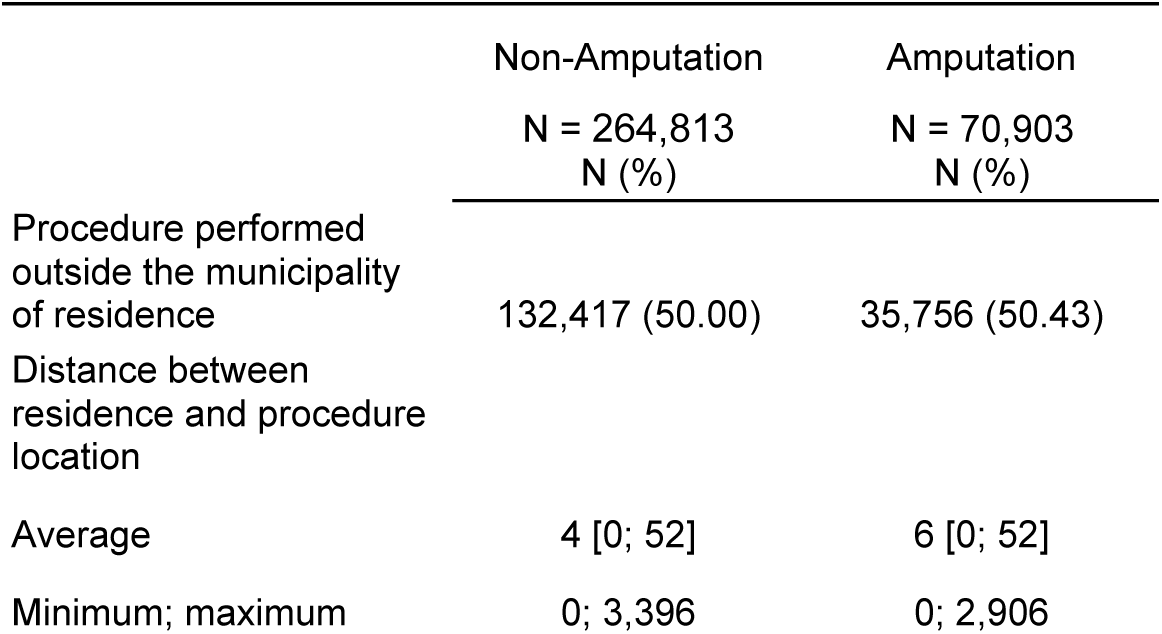
Analysis of Procedures and Care Location in Relation to Residence.

|  | Non-Amputation<br>N = 264,813<br>N (%) | Amputation<br>N = 70,903<br>N (%) |
| --- | --- | --- |
| Procedure performed outside the municipality of residence | 132,417 (50.00) | 35,756 (50.43) |
| Distance between residence and procedure location |  |  |
| Average | 4 [0; 52] | 6 [0; 52] |
| Minimum; maximum | 0; 3,396 | 0; 2,906 |

Table 5 presents factors associated with amputation risk (adjusted odds ratios). Having procedures outside the municipality of residence slightly raise odds (OR 1.017; p = 0.0442). Distance per 100 km has a minimal non-significant effect (OR 0.992; p = 0.0724). Females have significantly lower odds than males (OR 0.749; p < 0.0001). Compared to the Southeast region, the Midwest shows higher odds (OR 1.283), the Northwest slightly higher (OR 1.112), while the North (OR 0.756) and South (OR 0.863) show lower odds (all p < 0.0001). Fully state-managed hospitals show higher odds than fully municipal-managed ones (OR 1.098; p < 0.0001).

**Table 5.**
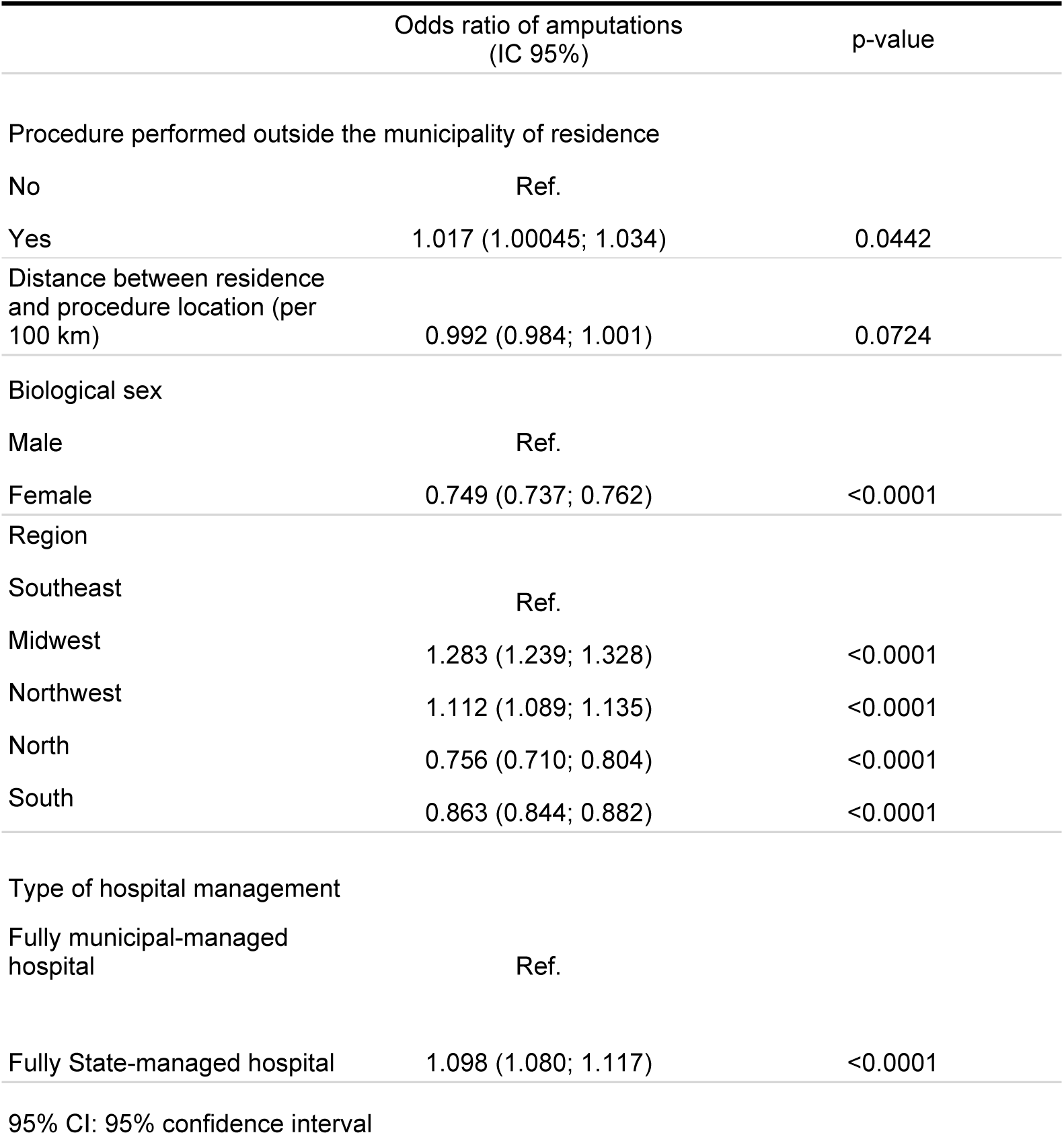
Univariate logistic regression models for amputation by Procedure, Distance, Sex, Region and Hospital Management.

A total of 17,900 deaths were recorded in the study population, including 6,673 among patients who underwent amputation and 11,227 among those without amputation. Overall, mortality accounted for 5.3% of all hospitalizations, with a higher frequency among amputated patients (9.4%) compared with non-amputated individuals (4.2%).

Among patients who underwent amputation, those who died were generally older (mean age 74 years) compared with survivors (mean age 67 years). Female patients represented a higher proportion of deaths (47.6%) than survivors (36.9%). Most deaths occurred in hospitals under municipal full management, and 52.8% of fatal cases required intensive care unit (ICU) admission.

Mortality rates showed modest fluctuations over the study period, with a gradual decline from 2015 to 2024. Regional differences were observed: the Southeast accounted for the largest proportion of cases, while higher mortality proportions were observed in the Central-West and North regions.

Table 6 shows logistic regression results for mortality in 70,903 amputation cases. Female sex increases odds (OR 1.557). Each 10-year increase in age raises the risk by 54% (OR 1.540). State-managed hospitals show slightly lower mortality than municipal ones (OR 0.900). ICU admission greatly increases the risk (OR 6.867). Procedures performed outside the patient’s municipality reduce the odds (OR 0.912), and each 100 km of travel distance also slightly lowers the risk (OR 0.963). Compared to the Southeast region, the Central-West (OR 1.193) and North (OR 1.293) have higher mortality, while the South (OR 0.847) has lower. Year by year, mortality risk declines modestly (OR = 0.956).

**Table 6:** Simple logistic regression models for mortality among 70,903 amputation cases.

|  | Odds Ratio (CI<br>95%) | p-value |
| --- | --- | --- |
| Patient sex |  |  |
| Male | Ref. |  |
| Female | 1.557 (1.480;<br>1.638) | <0.0001 |
| Age (per 10 years) | 1.540 (1.506;<br>1.574) | <0.0001 |
| Type of hospital management |  |  |
| Municipal full management | Ref. |  |
| State full management | 0.900 (0.856;<br>0.947) | <0.0001 |
| ICU |  |  |
| No | Ref. |  |
| Yes | 6.867 (6.513;<br>7.241) | <0.0001 |
| Procedure performed outside the municipality of residence |  |  |
| No | Ref. |  |
| Yes | 0.912 (0.867;<br>0.959) | 3 |
| Distance between residence and procedure location (per 100 km) | 0.963 (0.934;<br>0.991) | 123 |
| Region |  |  |
| Southeast | Ref. |  |
| Central-West | 1.193 (1.083;<br>1.313) | 3 |
| Northeast | 1.041 (0.979;<br>1.107) | 1.946 |
| North | 1.293 (1.080;<br>1.537) | 43 |
| South | 0.847 (0.788;<br>0.909) | <0.0001 |
| Year | 0.956 (0.947;<br>0.964) | <0.0001 |
| 95% CI: 95% confidence interval |  |  |

Length of hospital stay ranged from 0 to 344 days, with 97.6% of admissions lasting ≤30 days. Only 2.4% exceeded this duration. Median hospital stay was 5 days for men and 6 days for women. Regional medians varied modestly, ranging from 5 days in the Southeast and South to 7 days in the Central-West.

Table 7 presents regression models for hospital stay among 70,903 amputation cases. Female sex shows a slight increase in odds of longer stay (OR 1.039). Age per 10 years isn’t significant (OR 0.997). State-managed hospitals have modestly lower odds than municipal hospitals (OR 0.974). Procedures outside the patient’s municipality (OR 0.903) and every 100 km increase in distance (OR 0.982) lower odds of longer stay. Compared with the Southeast, the Central-West (OR 1.095) and the North (OR 1.120) have higher stays, while the South has lower stays (OR 0.857). Over time, stay length decreases modestly (OR 0.991 per year).

**Table 7.** Simple regression models for length of hospital stay among the 70,903 amputation cases.

|  | Odds Ratio (CI<br>95%) | p-value |
| --- | --- | --- |
| Patient sex |  |  |
| Male | Ref. |  |
| Female | 1.039 (1.023;<br>1.055) | <0.0001 |
| Age (per 10 years) | 0.997 (0.992;<br>1.003) | 3.420 |
| Type of hospital management |  |  |
| Municipal full management | Ref. |  |
| State full management | 0.974 (0.960;<br>0.989) | 9 |
| Procedure performed outside the municipality of residence |  |  |
| No | Ref. |  |
| Yes | 0.903 (0.888;<br>0.917) | <0.0001 |
| Distance between residence and procedure location (per 100 km) | 0.982 (0.975;<br>0.988) | <0.0001 |
| Region |  |  |
| Southeast | Ref. |  |
| Central-West | 1.095 (1.047;<br>1.145) | 1 |
| Northeast | 0.973 (0.956;<br>0.989) | 14 |
| North | 1.120 (1.064;<br>1.180) | <0.0001 |
| South | 0.857 (0.842;<br>0.873) | <0.0001 |
| Year | 2.694 (0.991;<br>0.991) | <0.0001 |
| 95% CI: 95% confidence interval |  |  |

## Discussion

This retrospective, population-based study encompassed most of the Brazilian population, given that over 72% of citizens rely on the public healthcare system, providing the strength of a large-scale epidemiologic analysis using national, mandatory administrative databases. Approximately 335,000 cases of peripheral arterial disease (PAD) were analyzed, with more than 70,000 amputations recorded among these patients over ten years. This represents the largest national study to date on the subject in Brazil, a country of continental dimensions. Data were obtained from DATASUS, the national public health database, which includes information exclusively from the public sector, thereby excluding patients receiving private care or those paying out of pocket.

The focus on ICD codes for atherosclerosis and arterial thrombosis ensures consistent case identification. At the same time, the use of geographic distance as determined by healthcare providers provides valuable insight into the systemic inequalities that influence amputation rates. The design effectively connects the spatial and demographic dimensions of vascular care delivery to major outcomes, such as amputation, mortality, and hospitalization length, making it a powerful model for health policy evaluation across almost the entire middle-income country.

Clinical management accounted for about half of all interventions, confirming the predominance of conservative or non-invasive approaches in PAD treatment within the public system.(16) The persistently high number of amputations, however, reflects the advanced stage at which many patients reach specialized care, often after failed conservative or endovascular attempts. Prior studies in both Brazil and high-income settings have demonstrated that late diagnosis, limited access to vascular specialists, and delayed revascularization substantially increase the likelihood of major limb loss. (17–19) These findings reinforce that amputation rates are not merely a reflection of disease severity but also an indicator of systemic inefficiency in the organization and delivery of vascular services.

Patients who underwent amputation presented a more severe clinical profile, with higher rates of emergency admission and ICU demand, consistent with advanced ischemia, infection, or treatment failure. These clinical characteristics suggest that most amputations occurred in acute or critical settings rather than as planned procedures, underscoring the lack of early detection and multidisciplinary management. Although overall access to care—measured by municipal distance and management type—appeared similar across groups, outcomes differed markedly, suggesting that organizational quality and the timing of intervention may play a larger role than structural access alone. (13)

Sex-specific differences were also an important finding. Female patients had a significantly lower likelihood of undergoing amputation than males, aligning with prior evidence that PAD tends to present differently across sexes, with men often developing more extensive atherosclerotic lesions or presenting later in the disease course. However, among those who required amputation, women exhibited higher postoperative mortality, and longer hospital stays compared to men. These disparities may reflect on vessel caliber, underrepresentation of women in vascular clinical trials, or delays in diagnosis and referral, which together can worsen outcomes despite a lower overall incidence of limb loss. (19,20) Further investigation into gender-specific disease presentation and treatment response is essential to optimize PAD management in women.

Amputation rates also varied markedly across Brazilian regions, reflecting the uneven distribution of vascular care infrastructure. The Southeast and Northeast accounted for the largest absolute numbers of procedures, consistent with their population size and hospital density. Yet, even in the Southeast—which concentrates the greatest number of vascular surgeons—surgeon-to-population ratios remain insufficient compared to demand. (21) This imbalance likely contributes to procedural delays, reduced opportunities for limb salvage, and consequently, higher amputation incidence. Similar patterns have been reported in other middle-income countries, where geographic and socioeconomic inequalities strongly determine vascular outcomes. (22,23) Expanding regional referral networks and ensuring timely revascularization could substantially reduce preventable amputations nationwide.

Over time, a modest upward trend in amputation rates was observed. Comparable temporal patterns have been described internationally, with some regions reporting declines following the widespread adoption of endovascular techniques, while others experience recurrent increases due to population aging and diabetes prevalence. (23–25) The Brazilian trend may reflect both demographic transition and systemic strain in vascular care capacity. (21) Sustained investment in early screening, foot care programs, early clinical treatment and local revascularization capability remains key to reversing this trajectory.

Regression models confirmed that demographic, structural, and clinical variables jointly influenced amputation risk. Older age and treatment outside the home municipality were associated with increased odds of limb loss, suggesting that both biological vulnerability and geographic barriers remain relevant. Procedures performed in state-managed hospitals carried higher amputation odds than those in municipally managed centers, implying that administrative and resource allocation differences may directly impact outcomes. Structural determinants—such as hospital type, staffing, and referral dynamics—have similarly been linked to PAD prognosis in other settings. (19)

The high mortality observed among patients undergoing amputation aligns with extensive international literature demonstrating the poor long-term prognosis of this population. Meta-analyses report mortality rates exceeding 50% within two years of non-traumatic amputation. (26,27) In Brazil, comparable data show 30-day mortality of 27% and two-year mortality reaching 77% among elderly patients with vascular or infectious etiologies. (28) These findings position amputation not only as a local therapeutic endpoint but as a sentinel event signaling systemic disease burden and frailty. Multidisciplinary prevention strategies—including early revascularization, optimization of diabetes and renal disease, and structured rehabilitation—are therefore critical to improve survival after amputation.

Regarding hospitalization time, the relatively short stays observed contrast with longer durations reported in high-income nations such as Denmark. (29) This discrepancy may reflect institutional efficiency or, conversely, premature discharge driven by limited bed capacity. The regression analysis also demonstrated a significant temporal effect, with mean hospital stay increasing by a factor of 2.694 after 2015, suggesting evolving clinical or administrative practices that warrant closer examination. Shorter hospitalizations can reduce costs and hospital risks but must be balanced against adequate postoperative care and follow-up to prevent complications and readmissions.

Regional disparities in outcomes following amputation likely mirror broader inequalities in healthcare access, infrastructure, and specialist distribution across Brazil. Patients from the North and Northeast face substantial barriers to specialized vascular care, often presenting with advanced disease and higher amputation risk. In contrast, the Southeast and South benefit from denser hospital networks and greater capacity for early intervention. (21) Addressing these structural inequities—through improved referral systems, expansion of multidisciplinary foot care programs, and equitable resource allocation—represents a crucial step toward aligning Brazilian PAD outcomes with international standards and reducing the national burden of preventable limb loss and mortality.

### Limitations

This study has inherent limitations to its retrospective, cross-sectional design, which precludes causal inferences and restricts the findings to associations and temporal trends. Because DATASUS data are anonymized and hospitalization-based, analyses were performed per admission rather than per patient. As a result, it was not possible to determine whether the same individual underwent multiple procedures over time—such as sequential clinical treatment, revascularization, and subsequent amputation—which could have provided insight into treatment failures or disease progression.

In addition, the database lacks individual-level clinical and demographic variables, including comorbidities, cardiovascular risk factors (such as smoking, diabetes, hypertension, obesity, and physical inactivity), and family or personal history of PAD. The absence of these variables limits the ability to adjust confounders and to identify specific risk factors associated with amputation or mortality. Finally, reliance on secondary administrative records may introduce inconsistencies in data entry or coding, and the lack of longitudinal tracking prevented evaluation of long-term outcomes.

### Conclusion

Geographic and organizational disparities significantly influence amputation outcomes among patients with peripheral arterial disease treated in Brazil’s public healthcare system. Greater travel distance and treatment outside the municipality of residence are associated with increased amputation risk, underscoring the importance of timely access to specialized vascular care. Women have a lower likelihood of amputation; however, when amputation occurs, they experience higher mortality and longer hospitalization. Overall mortality among amputees is elevated, and variations in hospital stay reflect differences in severity and resource availability across regions.

